# How do clinician and parent reported data differ? An analysis of similarity and difference in the datasets from a cross-syndrome genetics cohort study(GenROC)

**DOI:** 10.1101/2025.09.03.25334983

**Authors:** KJ Low, H Day, GenROC Consortium, M Thanthilla, C Davis, HV Firth, CF Wright

**Author notes:** Corresponding Author: K J Low.

## Abstract

**Background:** Parent/patient-reported datasets provide ready access to phenotypic data for monogenic neurodevelopmental disorders yet their concordance with clinical data is unclear.

**Methods:** In the GenROC study 547 children (mean age 7.6y, balanced sex ratio) had parallel parent-reported(PRD) web questionnaires and clinician-reported (CRD) Human Phenotype Ontology(HPO) proformas. We compared the two sources per participant by system, gene and gene group and overall for quantity, detail and similarity.

**Results:** 547 probands were analysed ranging in age from infancy to 16 years (mean 7.6) with similar gender distribution. PRD provided more terms for dental, gastroenterology, immunology and respiratory systems and for vision, (p <0.001 for all) and to a lesser degree for cardiac (p=0.0012). CRD provides more detail than PRD for most gene subgroups, combined systems and for neurology(p<0.001). Similarity scores were low overall per participant(mean 0.38 for combined) . Similarity scores were highest for cardiac (mean 0.74) and lowest for ENT(mean 0.34) There was minimal difference in similarity scores across gene groups or between the top 10 genes -scaffold adaptor gene groups had the highest (mean 0.43) as did *STXBP1*(mean 0.5) and *CACNA1A*(0.49). CRD is more similar to published syndrome phenotypes for syndromic genes.

**Conclusions:** Parents reported more common childhood phenotypes, such as asthma and dental issues, whilst clinicians provided clinical phenotype descriptors, such as brain morphology and seizure semiology. It is important to understand the differences when designing studies and utilising datasets to appreciate their strengths and limitations.

**What is already known on this topic:** Parent-reported data are increasingly used in rare disease research due to their accessibility and breadth. Previous studies have shown that such data can be consistent with published literature, particularly in syndromic conditions. However, direct comparisons between parent-reported and clinician-reported data at the individual level have been limited, leaving a gap in understanding the reliability and granularity of these data sources.

**What this study adds:** This study provides the first large-scale, individual-level comparison of parent-reported and clinician-reported phenotypic data across a cross-syndrome cohort. It demonstrates that while both sources contribute similar quantities of data, they differ in content and detail. Parents tend to report common childhood and lived experience phenotypes, whereas clinicians provide more specific clinical descriptors. The study also shows that clinician data are more consistent with published syndrome phenotypes, especially in syndromic genes.

**How this study might affect research, practice or policy:** These findings highlight the complementary nature of parent and clinician data in rare disease research. Future studies and registries should consider integrating both sources to enhance phenotypic richness and accuracy. Policymakers and researchers designing data collection tools or machine learning applications should account for the strengths and limitations of each data type, ensuring that lived experience data are not overlooked in phenotype descriptions.

## Introduction

Data are limited regarding phenotypes and natural history in rare monogenic neurodevelopmental disorders (NDDs) (1, 2). This data gap has led to many parent/patient-led foundations, as well as clinicians and researchers setting up parent/patient-reported natural history studies, some of which are run independently and many of which are hosted within wider platforms such as RareX and Simon’s searchlight(3, 4). Due to the obstacles and barriers to rare disease research these studies provide important accessible data for researchers and clinicians (5).

A scoping review identified six studies(6–11) in monogenic neurodevelopmental disorders which had included any comparisons of data provided from parents, clinicians and other sources. Three of the reports were derived from one overall study based on a cohort of individuals with chromosome 6 aberrations, and data derived from a web-based survey from parents(7). Parent-reported data were compared to the published literature. In one of their studies, they also made a comparison of parent-derived data for 20 individuals compared to their medical records(7). The results focussed on data consistency between parent-reported data and phenotypic themes derived from the published literature. Another study reported consistency of parent reported data in Simons Searchlight for *SLC6A1* compared with the published literature(9). This study has subsequently been cited by five publications regarding *SLC6A1* demonstrating the potential reach and impact on the literature of parent-reported data. Two further studies report on GenIDA collated parent-reported data in Koolen de Vries syndrome and *DDX3X* and briefly report on data consistency with published phenotypes.(10, 11)

Given the drive towards parent-reported online survey data as the primary phenotype and natural history data source for NDDs, we sought to investigate the consistency, similarity and differences in granularity of data provided by parents and clinicians on an individual patient level. We used a cross-syndrome cohort in order to better inform understanding of and decision-making regarding data sources and their applications in NDDs in the future.

## Methods

Data were derived from the GenROC study, a UK based cross syndrome cohort study of children with NDDs under 16 years of age. All participants had a confirmed pathogenic or likely pathogenic change in a single gene identified through clinical testing or through another research study. The full protocol for the study is published elsewhere(12). The GenROC study received Research Ethics Committee (REC) approval on 15 December 2022 and Health Research Authority approval on 9 February 2023.

Parents were asked to complete an online web-based survey in REDCap electronic data capture tools hosted at the University of Bristol. REDCap (Research Electronic Data Capture) is a secure, web-based software platform designed to support data capture for research studies, providing 1) an intuitive interface for validated data capture; 2) audit trails for tracking data manipulation and export procedures; 3) automated export procedures for seamless data downloads to common statistical packages; and 4) procedures for data integration and interoperability with external sources(13,14). The survey included a broad variety of questions regarding their child’s development, growth and medical features. Some questions were checkbox choices but for each physical system parents were asked to complete a free-text box. They were given the option of leaving the box blank if not relevant for their child. Parent free-text responses for the cohort were then separated by clinical system to avoid later inferences from the curator across the systems. Three non-specialist curators (two medical students and one junior doctor) then reviewed the free-text data and curated them into Human Phenotype Ontology (HPO) terms as per the April 2024 version of hpo.jax.org.

For each participant in GenROC, their responsible clinical geneticist (or delegate) was invited to complete a clinical proforma via a web-based survey. This included genomic variant data alongside growth, prenatal and phenotype terms grouped by the same body systems as in the parent survey. Clinicians were requested to provide HPO terms but were given the option of free text response. Where free text response was provided this was curated into HPO terms by expert curation (a clinical geneticist). These curated data were then processed in Python using the pyhop.ontology package (https://pyhpo.readthedocs.io/en/stable/). All the functions used (but none of the notebooks) can be found here: https://github.com/JGIBristol/GenROC_Public.

HPO terms from the clinician and parent were then analysed on an individual participant level using the following approaches. Probands where data were only collected from one source were excluded from this analysis.

For the purposes of analyses below we have grouped genes into gene group categories. The categories were defined using the following process. The gene list was first uploaded into PantherDB(15,16) which provides groupings of genes based on molecular function.

After this initial step unclassified genes were manually reviewed with gene categories. Categories were expanded to include further genes based on molecular and clinical characteristics. (See Table 1 for list of genes and curated categories).

**Table 1:**
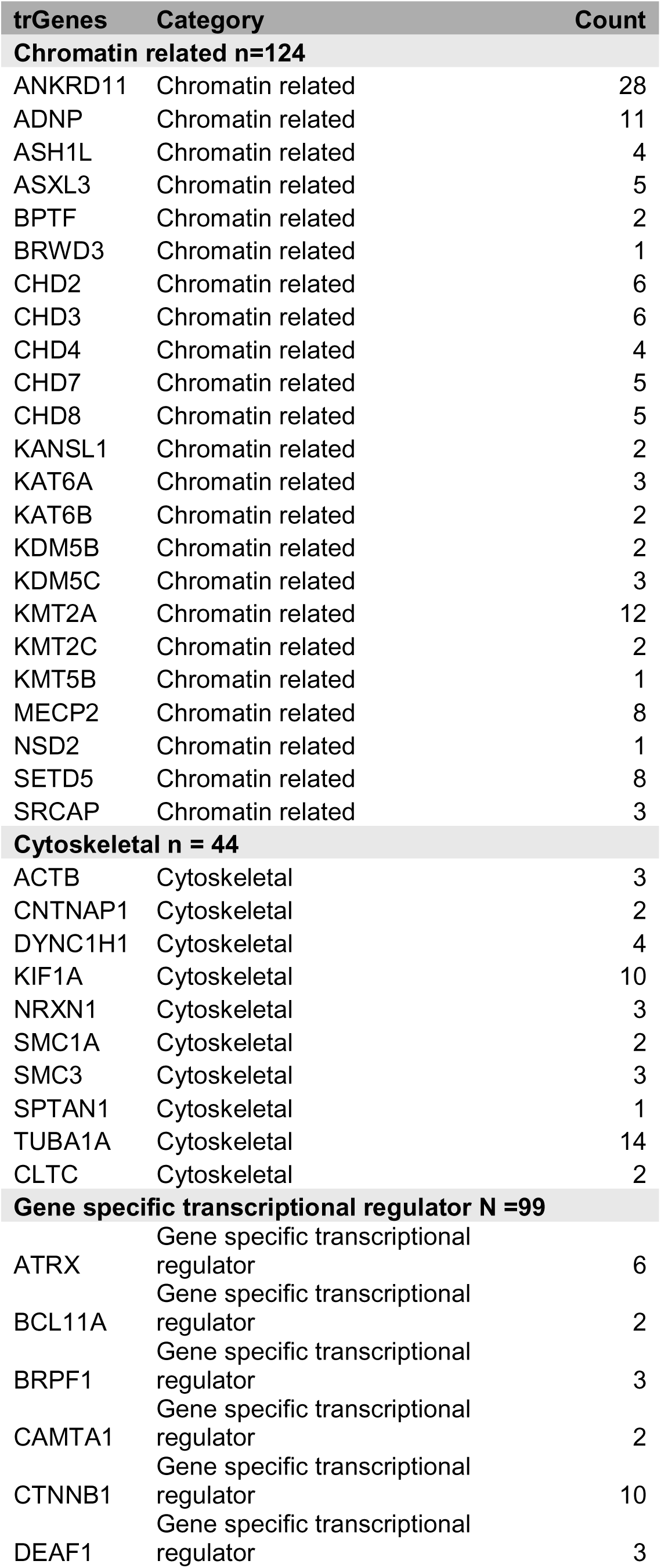
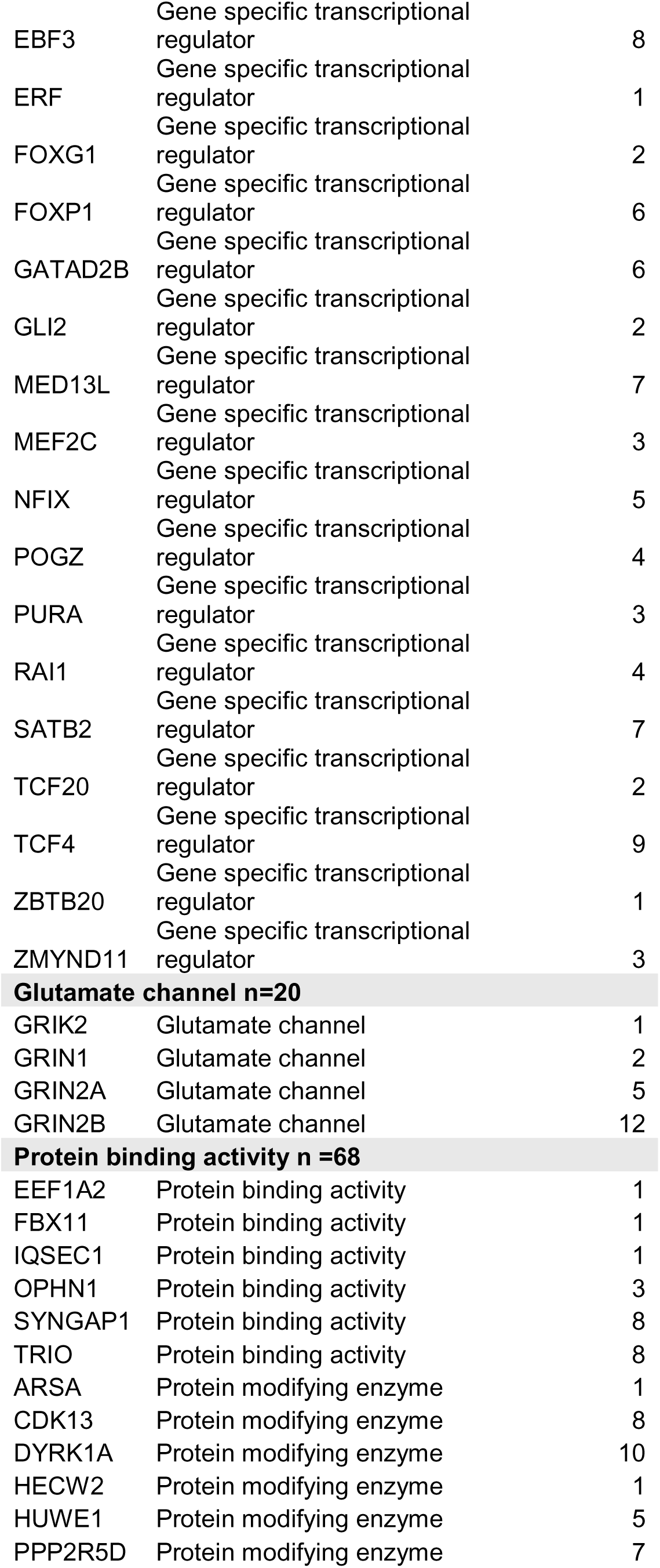
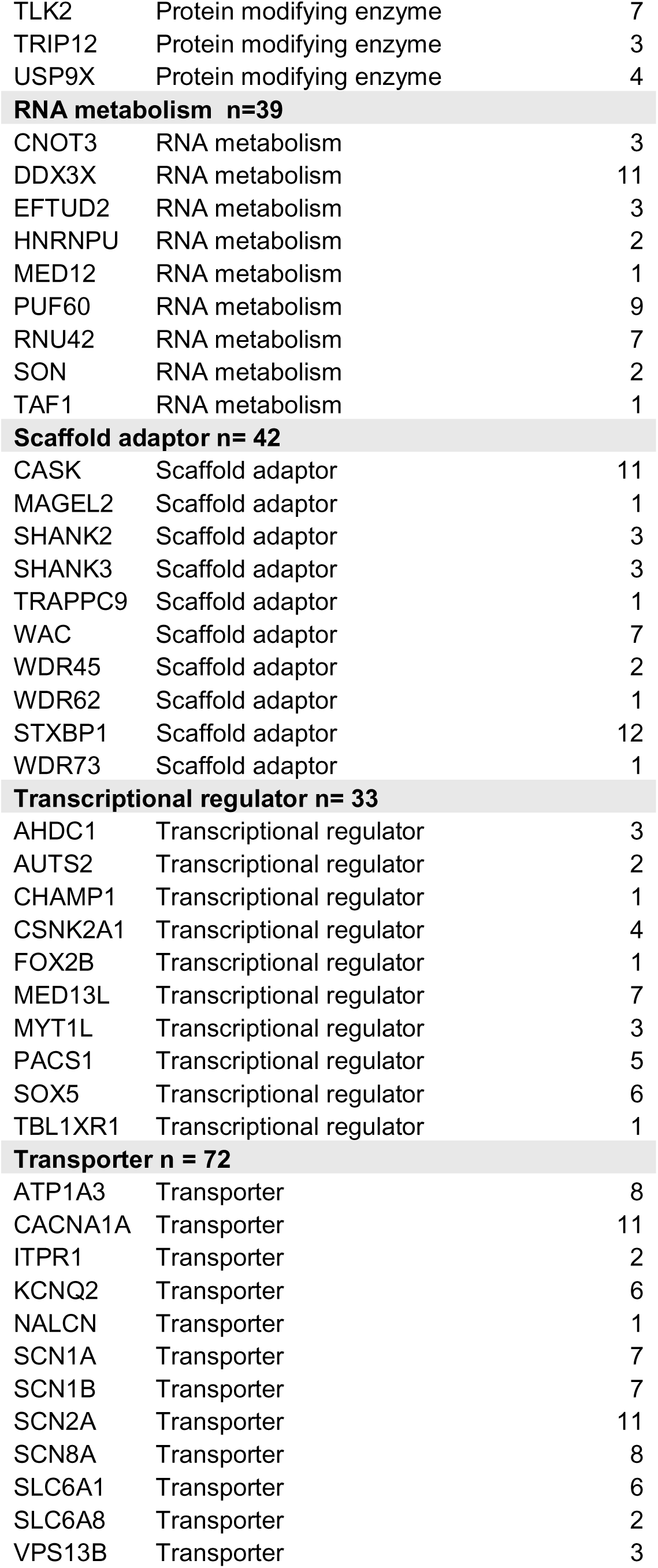
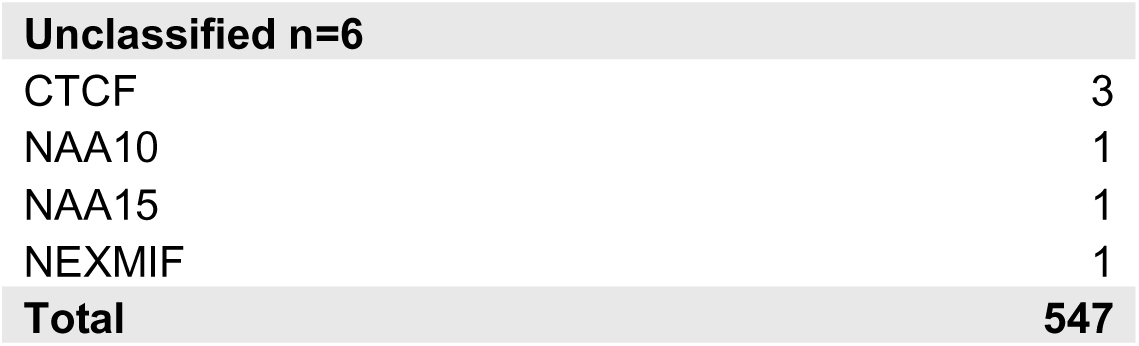
Table of genes groups and genes in GenROC including number of participants recruited.

| trGenes | Category | Count |
| --- | --- | --- |
| **Chromatin related n=124** |  |  |
| ANKRD11 | Chromatin related | 28 |
| ADNP | Chromatin related | 11 |
| ASH1L | Chromatin related | 4 |
| ASXL3 | Chromatin related | 5 |
| BPTF | Chromatin related | 2 |
| BRWD3 | Chromatin related | 1 |
| CHD2 | Chromatin related | 6 |
| CHD3 | Chromatin related | 6 |
| CHD4 | Chromatin related | 4 |
| CHD7 | Chromatin related | 5 |
| CHD8 | Chromatin related | 5 |
| KANSL1 | Chromatin related | 2 |
| KAT6A | Chromatin related | 3 |
| KAT6B | Chromatin related | 2 |
| KDM5B | Chromatin related | 2 |
| KDM5C | Chromatin related | 3 |
| KMT2A | Chromatin related | 12 |
| KMT2C | Chromatin related | 2 |
| KMT5B | Chromatin related | 1 |
| MECP2 | Chromatin related | 8 |
| NSD2 | Chromatin related | 1 |
| SETD5 | Chromatin related | 8 |
| SRCAP | Chromatin related | 3 |
| **Cytoskeletal n = 44** |  |  |
| ACTB | Cytoskeletal | 3 |
| CNTNAP1 | Cytoskeletal | 2 |
| DYNC1H1 | Cytoskeletal | 4 |
| KIF1A | Cytoskeletal | 10 |
| NRXN1 | Cytoskeletal | 3 |
| SMC1A | Cytoskeletal | 2 |
| SMC3 | Cytoskeletal | 3 |
| SPTAN1 | Cytoskeletal | 1 |
| TUBA1A | Cytoskeletal | 14 |
| CLTC | Cytoskeletal | 2 |
| **Gene specific transcriptional regulator N =99** |  |  |
| ATRX | Gene specific transcriptional regulator | 6 |
| BCL11A | Gene specific transcriptional regulator | 2 |
| BRPF1 | Gene specific transcriptional regulator | 3 |
| CAMTA1 | Gene specific transcriptional regulator | 2 |
| CTNNB1 | Gene specific transcriptional regulator | 10 |
| DEAF1 | Gene specific transcriptional regulator | 3 |
| EBF3 | Gene specific transcriptional regulator | 8 |
| ERF | Gene specific transcriptional regulator | 1 |
| FOXG1 | Gene specific transcriptional regulator | 2 |
| FOXP1 | Gene specific transcriptional regulator | 6 |
| GATAD2B | Gene specific transcriptional regulator | 6 |
| GLI2 | Gene specific transcriptional regulator | 2 |
| MED13L | Gene specific transcriptional regulator | 7 |
| MEF2C | Gene specific transcriptional regulator | 3 |
| NFIX | Gene specific transcriptional regulator | 5 |
| POGZ | Gene specific transcriptional regulator | 4 |
| PURA | Gene specific transcriptional regulator | 3 |
| RAI1 | Gene specific transcriptional regulator | 4 |
| SATB2 | Gene specific transcriptional regulator | 7 |
| TCF20 | Gene specific transcriptional regulator | 2 |
| TCF4 | Gene specific transcriptional regulator | 9 |
| ZBTB20 | Gene specific transcriptional regulator | 1 |
| ZMYND11 | Gene specific transcriptional regulator | 3 |
| **Glutamate channel n=20** |  |  |
| GRIK2 | Glutamate channel | 1 |
| GRIN1 | Glutamate channel | 2 |
| GRIN2A | Glutamate channel | 5 |
| GRIN2B | Glutamate channel | 12 |
| **Protein binding activity n =68** |  |  |
| EEF1A2 | Protein binding activity | 1 |
| FBX11 | Protein binding activity | 1 |
| IQSEC1 | Protein binding activity | 1 |
| OPHN1 | Protein binding activity | 3 |
| SYNGAP1 | Protein binding activity | 8 |
| TRIO | Protein binding activity | 8 |
| ARSA | Protein modifying enzyme | 1 |
| CDK13 | Protein modifying enzyme | 8 |
| DYRK1A | Protein modifying enzyme | 10 |
| HECW2 | Protein modifying enzyme | 1 |
| HUWE1 | Protein modifying enzyme | 5 |
| PPP2R5D | Protein modifying enzyme | 7 |
| TLK2 | Protein modifying enzyme | 7 |
| TRIP12 | Protein modifying enzyme | 3 |
| USP9X | Protein modifying enzyme | 4 |
| **RNA metabolism n=39** |  |  |
| CNOT3 | RNA metabolism | 3 |
| DDX3X | RNA metabolism | 11 |
| EFTUD2 | RNA metabolism | 3 |
| HNRNPU | RNA metabolism | 2 |
| MED12 | RNA metabolism | 1 |
| PUF60 | RNA metabolism | 9 |
| RNU42 | RNA metabolism | 7 |
| SON | RNA metabolism | 2 |
| TAF1 | RNA metabolism | 1 |
| **Scaffold adaptor n= 42** |  |  |
| CASK | Scaffold adaptor | 11 |
| MAGEL2 | Scaffold adaptor | 1 |
| SHANK2 | Scaffold adaptor | 3 |
| SHANK3 | Scaffold adaptor | 3 |
| TRAPPC9 | Scaffold adaptor | 1 |
| WAC | Scaffold adaptor | 7 |
| WDR45 | Scaffold adaptor | 2 |
| WDR62 | Scaffold adaptor | 1 |
| STXBP1 | Scaffold adaptor | 12 |
| WDR73 | Scaffold adaptor | 1 |
| **Transcriptional regulator n= 33** |  |  |
| AHDC1 | Transcriptional regulator | 3 |
| AUTS2 | Transcriptional regulator | 2 |
| CHAMP1 | Transcriptional regulator | 1 |
| CSNK2A1 | Transcriptional regulator | 4 |
| FOX2B | Transcriptional regulator | 1 |
| MED13L | Transcriptional regulator | 7 |
| MYT1L | Transcriptional regulator | 3 |
| PACS1 | Transcriptional regulator | 5 |
| SOX5 | Transcriptional regulator | 6 |
| TBL1XR1 | Transcriptional regulator | 1 |
| **Transporter n = 72** |  |  |
| ATP1A3 | Transporter | 8 |
| CACNA1A | Transporter | 11 |
| ITPR1 | Transporter | 2 |
| KCNQ2 | Transporter | 6 |
| NALCN | Transporter | 1 |
| SCN1A | Transporter | 7 |
| SCN1B | Transporter | 7 |
| SCN2A | Transporter | 11 |
| SCN8A | Transporter | 8 |
| SLC6A1 | Transporter | 6 |
| SLC6A8 | Transporter | 2 |
| VPS13B | Transporter | 3 |

|  |  |
| --- | --- |
| **Unclassified n=6** |  |
| CTCF | 3 |
| NAA10 | 1 |
| NAA15 | 1 |
| NEXMIF | 1 |
| **Total** | **547** |

The following scores were generated for each proband for PRD and CRD:

### Quantity score

The number of HPO terms provided (high number = many HPO terms provided). We compared the CRD and PRD quantity result scores and counted the instances of “Parent more”, “Doctor more” and “Equal quantity” for System, gene category, and top 10 genes. For all 3 we calculated p values for “Doctor more” and “Parent more” using McNemar’s χ² test in R. Given the multiple comparisons, raw p values were adjusted by the Benjamini– Hochberg false-discovery-rate procedure(FDR).

### Detail score

A quantitative score reflecting distance in detail in the HPO branches between all possible pairs (one from PRD and one from CRD) of HPO codes for a given participant. For each pair of codes we assess if one code is an ancestor of another code in the HPO tree. If one is an ancestor of the other then the detail score increases for the source providing those data (PRD or CRD). The amount the score increases scales with the level of precision, e.g. if PRD says “Mild myopia HP:0025573” and CRD said “Myopia HP:0000545” then PRD would get a detail score increase of one. However, if the CRD had instead said “Abnormality of refraction HP:0000539” which is the direct ancestor of Myopia, then PRD detail score would increase by two. If not, the detail score does not change. If however, CRD has said “Nystagmus: HP:0000639” and PRD said ““Mild myopia HP:0025573” neither would score as these terms are unrelated. We compared the CRD and PRD detail results scores for “Doctor more” and “ Parent more” for System, gene category, and top 10 genes. P values were calculated using McNemar’s χ² test in R(3). Given the multiple comparisons, raw p values were adjusted by by the Benjamini–Hochberg false-discovery-rate procedure(FDR)(4).

Calculated Semantic Similarity scores: We used the similarity score system(13) for all pairs to assess similarity score by system and overall per participant. Pairwise semantic similarity scores ranging from 0 to 1 were calculated for gene pairs based on their associated HPO terms using the ontologySimilarity package in R (v4.2.1) (https://cran.r-/web/packages/ontologySimilarity/vignettes/ontologySimilarity-introduction.html). This method applies Lin’s information-theoretic approach to quantify term similarity within the HPO hierarchy, where a score of 1 indicates identical phenotypic annotations(18,19). A similarity score of 1 indicates an identical response. We manually reviewed 20 probands with the lowest similarity scores to assess for any common themes.

For the top 10 most recruited genes in GenROC we compared the PRD, CRD and combined HPO terms per participant with the associated HPO terms in G2P for each respective gene (https://www.ebi.ac.uk/gene2phenotype/ ) and calculated similarity scores. We first applied a Welch one-way ANOVA (20) in R to test global differences without assuming equal variances. Significant results were followed by all pairwise Welch t-tests, with Holm’s sequential adjustment(21).

Phenotype frequency by data source: we reviewed the numbers of HPO codes from PRD and CRD by system over the whole cohort. We reviewed which codes were most frequent and common to both sources and which were unique.

Finally, we manually reviewed the reporting of one common and important phenotype to determine reporting consistency and assess for differences. We selected epilepsy as this is frequent in the cohort and a phenotype that most parents would be aware of. For this analysis we searched all HPO terms in PRD and CRD for HP:0001250 or a child code. This outputted a present/absent variable for each proband. For all “absent” cases an expert clinician also reviewed the HPO terms to check that none had been missed that were consistent with epilepsy. For all “present” cases these data were then stratified into PRD reported; CRD reported; and “both” and manually by seizure classification where these data were available. Comparison was made across the three categories to assess for any differences.

## Results

547participants had been recruited to the GenROC study at the time of this analysis, of which 477 participants had PRD and CRD. The amount of missing data varied by system as for some systems, one may have provided data but not the other and we elected to exclude these from systems analyses in order to allow for direct comparisons. For the ‘combined’ systems analysis, we only included the 477 probands who had PRD and CRD for each system.

CRD were provided by clinical geneticists or a delegate whose proforma was then checked and approved by the clinician. 69% of parents in the cohort reported the highest level of parental education at degree level or above, with 24% having achieved school leaver qualifications of some sort. Only 3% reported having not achieved any educational qualifications.

### Quantity scores

PRD and CRD provide a similar quantity of data but with some notable differences by system and gene group.

PRD and CRD scores showed a similar normal distribution (Figure1a) with most providing 5 or 6 terms per participant. Not many participants had more than 16 terms provided by either source but when this did occur this was mostly by CRD.

PRD provided much more data for dental, gastroenterology, immunology and respiratory systems and for vision, (p <0.001 for all, figure 1b). This was also true for cardiac but to a lesser degree (p=0.0012). There was no evidence of a difference for endocrine, ear, nose and throat, or renal. The systems with the fewest HPO codes provided (shown in figure 1b by “No HPO codes”) were cardiac, endocrine, immune and renal.

**Figure 1:**
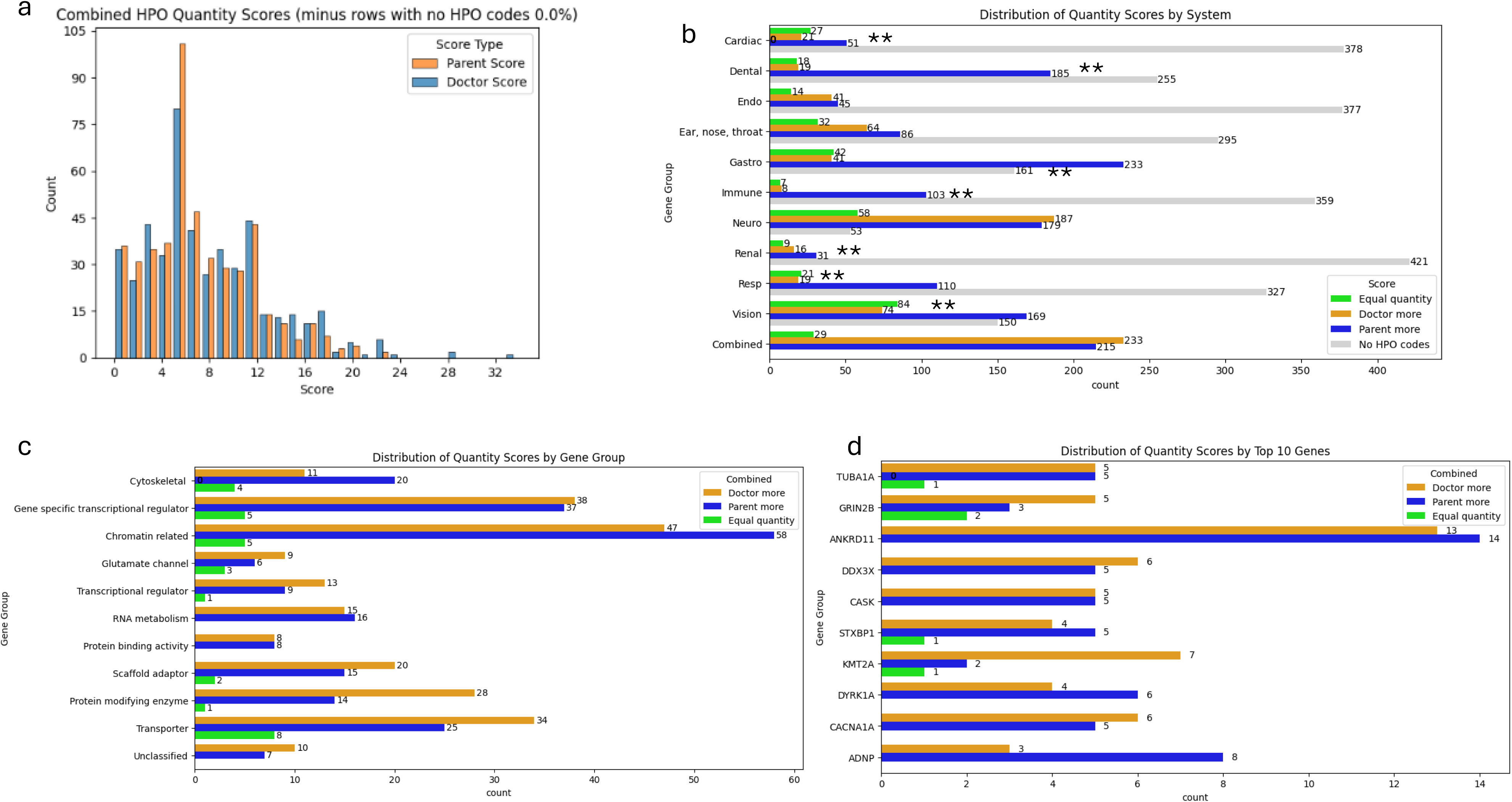
Quantity: a) Histogram of combined systems HPO quantity scores b) Distribution of quantity scores by system. c) Distribution of quantity scores by gene group d) Distribution of quantity scores for top 10 genes (highest recruited numbers in GenROC) ** = p value <0.05

For neurology approximately a third of the cohort had more terms provided by PRD and another third had more terms provided by CRD.

Most of the gene groups showed similar distributions for PRD and CRD(figure 1c). There was no statistical evidence of a difference in scores by gene category or by single gene (top 10 genes). Cytoskeletal and Chromatin related groups showed higher scores for PRD than CRD in comparison to Transporter and Protein modifying Enzyme groups which scored higher for CRD than PRD, albeit to a lesser extent. There were no striking differences by top 10 genes(figure 1d).

### Detail scores

CRD provides more detail than PRD for most gene subgroups, combined systems and for neurology:

PRD and CRD showed similar detail by system(figure 2a). The most notable exception is for neurology in which 60 probands had a higher detail score for CRD (p<0.001, heatmap figure 2b). This was seen to a much lesser extent in gastroenterology with 11 probands having higher scores for CRD. Vision was different to the rest of the systems with a small almost identical proportion of both higher CRD and PRD scores (28 probands had higher details scores for PRD; 24 probands had higher scores for CRD). Combined systems showed evidence for a difference with CRD having higher detail scores than PRD (p <0.001).

**Figure 2:**
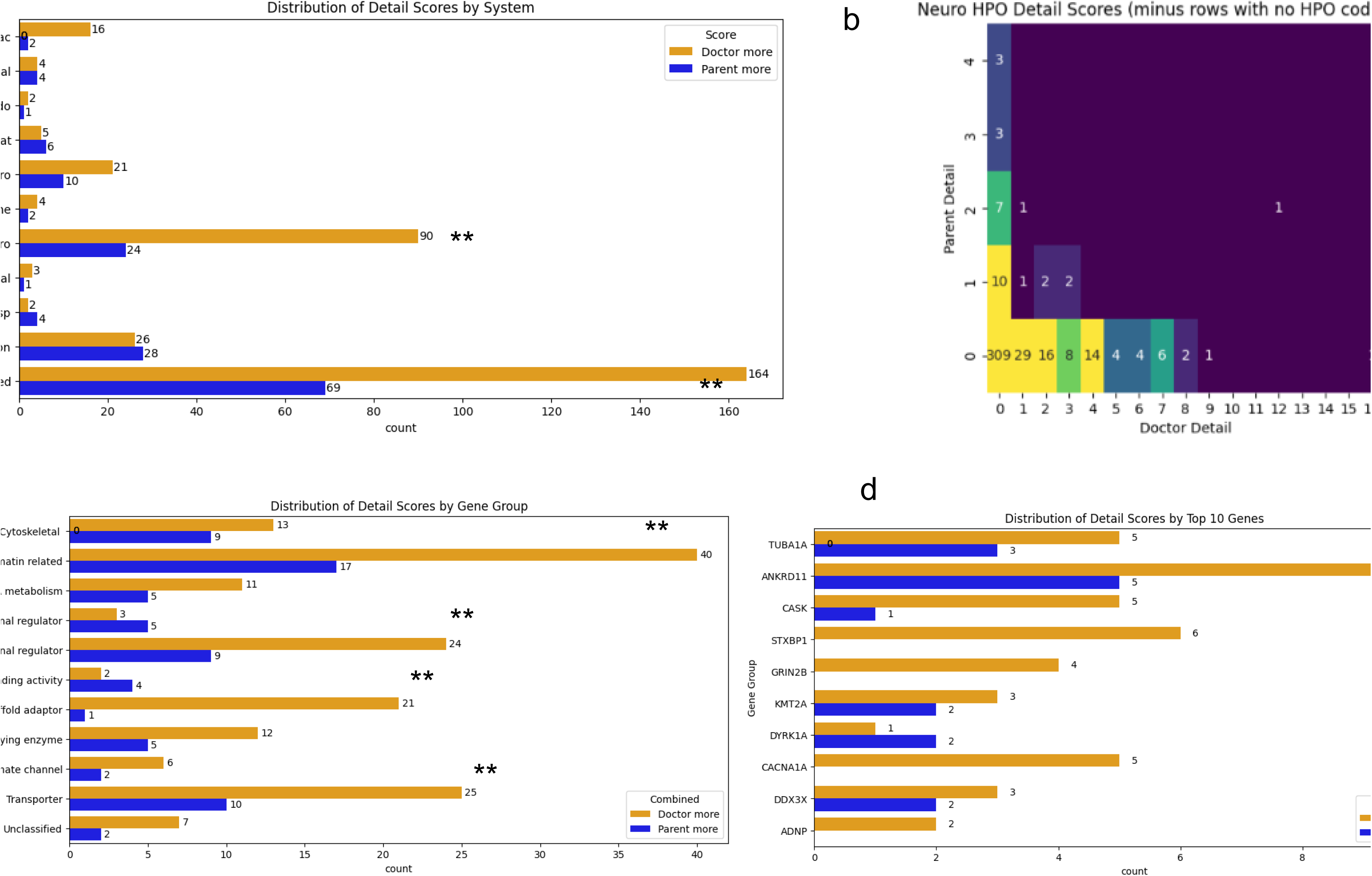
Detail: a) Distribution of detail scores by system. b) Heatmap of detail scores for neurology system c) Distribution of detail scores by gene group d) distribution of detail scores by top 10 recruited genes. ** = p <0.05

When looking just at the “Parent more” and “Doctor more” categories all but 2 of the gene subgoups scored higher for detail for CRD than PRD (figure 2c) and the evidence was strongest for this the Scaffold adaptor group (p<0.01) as well as for Chromatin related(p=0.035), Gene specific transcriptional regulator (p=0.045) and Transporter(p=0.045) groups. There was no strong evidence for a difference by top 10 gene(figure 2d).

### Similarity

Similarity scores were low overall per participant but differed somewhat by system,gene and gene group(figure 3). Doctors provided more unique terms for neurology compared to parents who provided more unique terms for gastro, dental and respiratory systems. CRD are more similar to published syndrome phenotypes.

**Figure 3:**
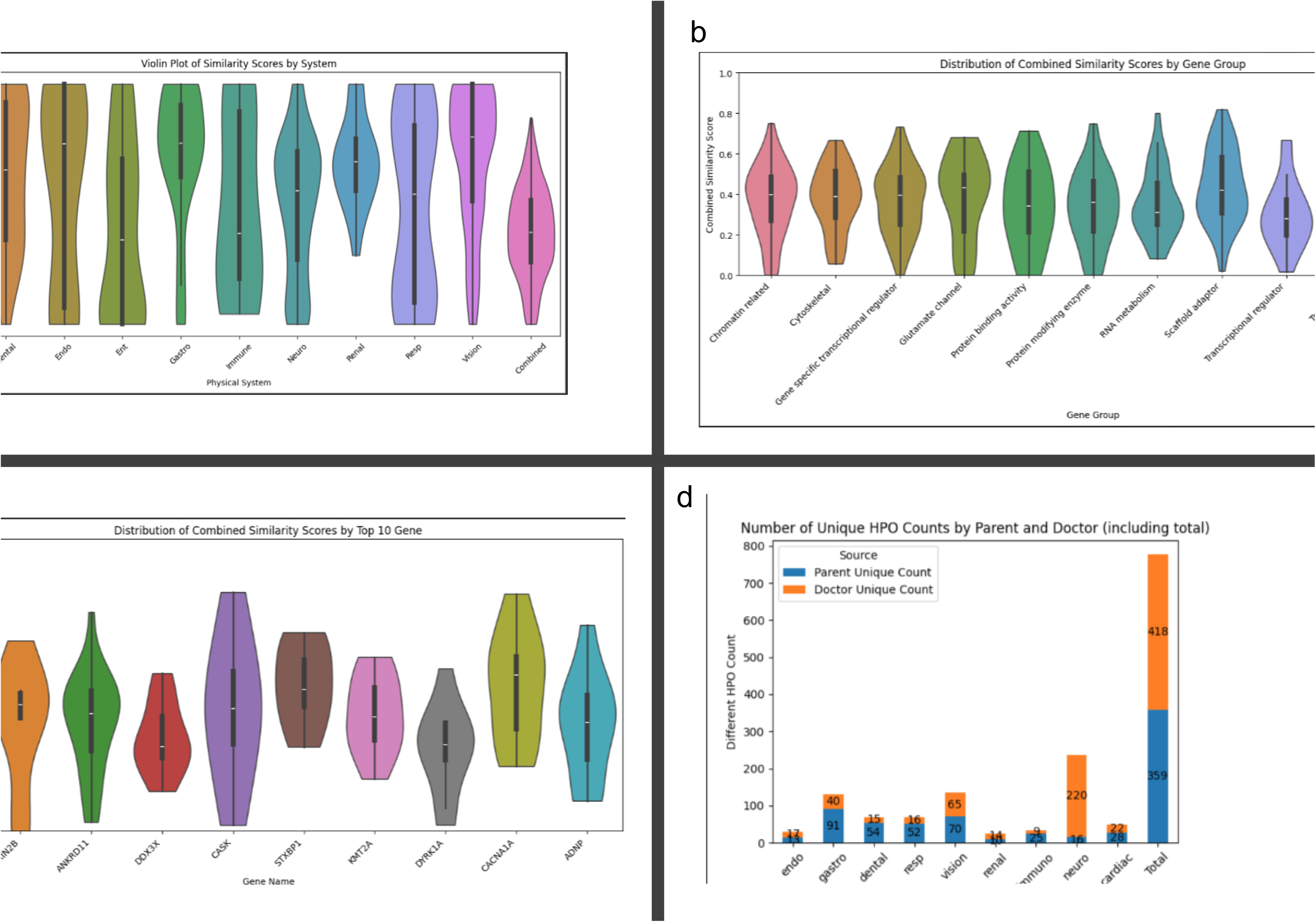
Similarity : a) Violin plot with box and whisker overlay depicting similarity score distribution for all systems and depicting median and upper and lower quartiles. b) Violin plot of similarity scores by gene groups c) Violin plot of similarity scores by top 10 recruited genes d) Number of unique HPO codes from PRD and CRD by system

The scores are highest for cardiac (mean 0.75; p <0.05) and followed by gastro(mean 0.70; p<0.05), vision(0.69;p<0.05) and dental(0.63;p<0.05), whilst the systems that had the lowest scores was ENT(mean 0.43;p=0.8). (figure 3a). The mean similarity score for the combined systems was 0.38. Similarity scores were consistent across gene groups with a mean ranging from 0.30 (transcription al regulators) to 0.43 in the scaffold adaptor group (figure 3b). Similarity scores were highest for *STXBP1*(mean 0.5) and *CACNA1A*(0.49) (figure 3c). There was no evidence of a statistical difference by gene or gene subgroup.

Manual review of the 20 lowest similarity score participants showed that PRD provided terms regarding respiratory, feeding and gastroenterology , dental and autonomic features whereas CRD describe dysmorphic features and clear specific medical phenotypes in greater detail.

When comparing PRD with the published HPO terms in DDG2P for each gene (figure 4a) we found that half (*ANKRD11,DDX3X, KMT2A, DYRK1A, ADNP*) had evidence for a reduction in similarity score(4d) compared to CRD(4b) or when combining datasets(4c). For all 5 of these conditions CRD is more similar to DDG2P than PRD (p<0.05). This is also true when comparing PRD and the combined datasets to DDG2P. There is no evidence for a difference between CRD and combined for any of the genes analysed. There was no evidence of a difference between PRD and CRD with DDG2P for *TUBA1A, GRIN2B, CASK, STXBP1 or CACNA1A*.

**Figure 4:**
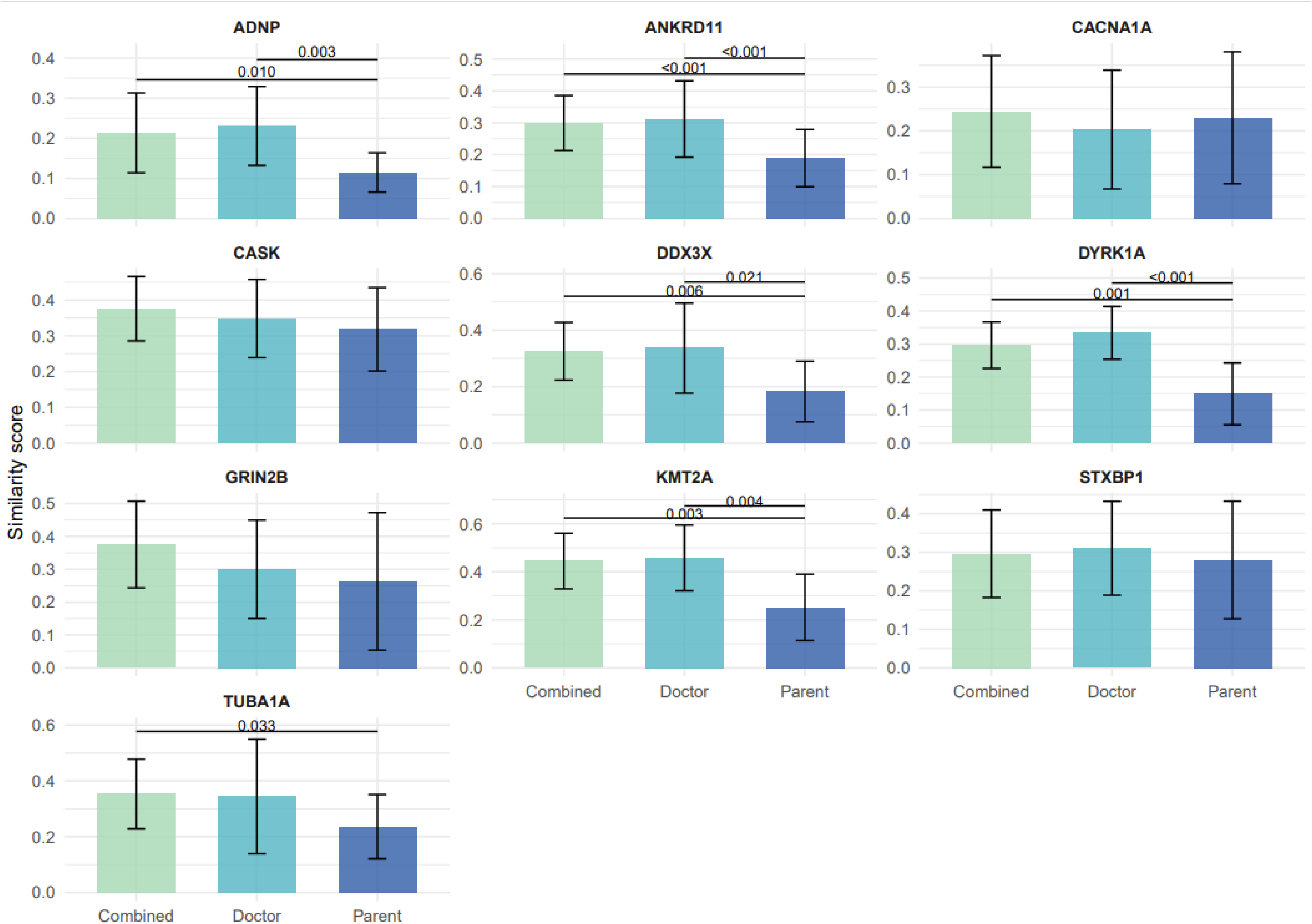
Similarity of reported data from parents, clinicians and combined with DDG2P published phenotypes. Clustered bar chart depicting similarity scores for each gene with DDG2P phenotypes for combined datasets, doctor only and parent only with error bars. P values are annotated where p <0.05 and the accompanying horizontal bars depicts the correlating comparison eg combined vs parent.

**Figure 5:**
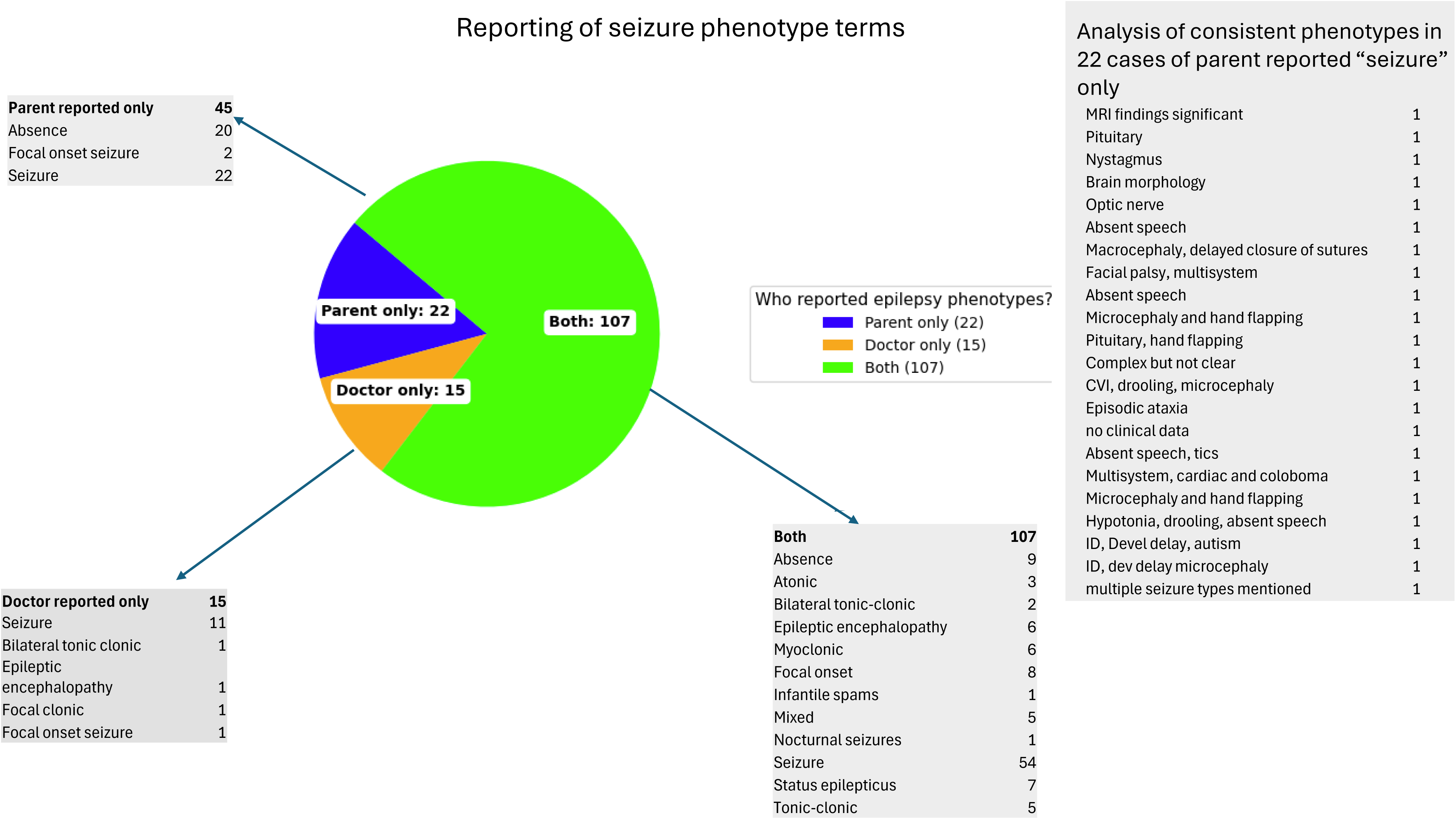
Analysis of seizure term reporting by PRD, CRD and both

**Phenotypic spread** analysis showed that CRD reported more terms with more detail in neurology but the most frequently reported terms are common to both datasets. When assessing the number of unique phenotypes provided there is a noticeable difference between systems with neurology alone contributing 220 of the 418 unique codes supplied by clinicians(figure 3e). Gastroenterology, dental and respiratory had more than double the number of unique codes supplied by PRD compared to CRD. Vision and cardiac had roughly equal numbers for both. Excluding developmental delay and intellectual disability, the 5 most frequently reported HPO terms in the combined dataset were identical to PRD top 5 and included hypotonia (198); constipation(127); seizure (119); strabismus (106) and Gastroesophageal reflux (85). CRD overlapped but also included Microcephaly (69) and hypertelorism (46).

Seizures were reported in 167 (35%) of the cohort(figure5). Of these, seizures were reported by both sources in 64%; by CRD only in 9% and in PRD only in 27%(Figure 4). For the 45 PRD-only cases, 20 had absence seizures, 2 had focal onset seizures and 22 just had the term “seizure”. Manual review of the 22 “seizure” cases showed significant neurological phenotypes which would be consistent clinically with coexistent seizures in all but 2 cases in which data were insufficient for analysis. Parents used predominantly medical terminology specifying “epilepsy” or “seizures” to describe the seizures with only two using “fits”. Two of the parents in the PRD only group used very specific terms “focal onset seizures”.

## Discussion

Rare disease registries have increased exponentially in the last decade with over 800 rare disease registries listed in a 2021 European report(22). However, whilst there may be an abundance, there have been concerns raised about the challenges of developing and sustaining a high-quality registry(23). However rare disease data are hard to obtain and collate due to the scarcity of patients, difficulty in locating them and wide geographical spread. Therefore, registry data represent a simple, quick and inexpensive way to undertake research. These sorts of data seem likely to be used in the future to create datasets for machine learning and other applications. It is therefore essential to understand the strengths and weaknesses of these data. A handful of previous studies have assessed parent-reported data with the published literature and concluded that they were reasonably consistent(6, 7, 9, 10, 11). However, to our knowledge no one has directly compared parent and clinician reported data for consistency. Our study sought to assess this consistency on an individual level across a large cohort of neurodevelopmental disorders to help inform the appropriate future use of such datasets.

We know from our GenROC qualitative study (25) that parents become experts in their child’s condition and spend a considerable amount of time and energy on educating professionals and directing and project managing their child’s care. They keep meticulous records. We also know from ThinkAloud(26) user testing of the GenROC parent proformas(12) that parents will look at their child’s clinic letters to help them complete the parent questionnaires. It is therefore to be expected that there should be relatively high data consistency between parents and clinicians, which is consistent with our results. We found particularly high similarity scores for the cardiac system, where most of the terms used were regarding cardiac malformations such as “ventriculo-septal defect (VSD)”. We would expect that these sorts of terms would be used in discussions with parents and in clinic letters and so would account for this level of consistency. The cardiac system also seems less likely to have as many lived experience phenotypes which might account for the similarity.

A strength of this study is that we have been able to undertake the analysis on an individual participant level comparing parent and clinician derived data. However, a limitation of the study is that there was high missingness that resulted in exclusions. However, this could simply reflect the nature of the disorders that were included in GenROC – by definition, these were neurodevelopmental genetic disorders and so many of these children may not have had particular problems in some of the systems (such as renal or immune) and so missing data may represent the lack of phenotype . The proportion of missing data for our analysis demonstrates the value of acquiring data from *both* parents and clinicians. By utilising this approach we were able to gather data across all areas in more individuals. By requesting data from two sources this maximised the number of individuals for whom we had at least some data provided rather than nothing at all.

Previous studies have tried to quantify the reliability of parent-reported data by comparing parent reported data with prevalence of phenotypes in the published literature.(9,10) These studies utilised a predefined generic GenIDA template in which parents were asked to respond “yes/no/I don’t know” to questions. This method allowed the authors to determine phenotypes in which a difference reached “statistical significance” such as movement disorder and hypotonia in the *DDX3X* cohort.(24) No significant differences were seen in the Koolen de Vries syndrome cohort(10). A strength of our study compared to these previous ones, is that parents were given free-text boxes which provided them with unlimited response options which could then be curated for comparison. Our analysis through similarity scores using the HPO ontology allows a more detailed analysis of consistency in reporting which may be important for granularity of phenotype description especially for future applications such as for machine learning datasets.

A possible limitation of our study is that we performed a curation step in order to convert free-text parent responses into HPO terms. In order to try and minimise introduction of clinician expertise into this step, we asked non-specialist medical students to do this work and also split the responses by system to ensure that the student could not make inferences across the dataset for an individual participant.

Another limitation of this study is the skewed nature of parental educational experience that is not representative of the general population.

Clinical proformas were completed by clinicial geneticists (or a delegate). These are busy clinicians who may have had very limited time to provide this data. We therefore noted differences between sites and clinicians with respect to the “fullness” of the clinician proforma. In addition, some sites used an appropriately trained research nurse, genetics trainee or genetic counsellor to complete the proforma. In those situations, they would also spend time consulting wider hospital records to ensure they were able to provide as comprehensive a set of answers as possible. This is unlikely to have been possible for the senior clinicians who had significant time restrictions due to competing clinicial priorities. As such it is likely that there could be a considerable difference in scores if it had been possible to perform sub-analyses based on data collectors. Nonetheless this reflects real-life data collection in clinical sites – there is likely to be variability in data reporting in any phenotyping study dependent on who is providing the data both in terms of experience, seniority and skill but also just as importantly in terms of allocated time for the task and motivation to do so.

Clinical geneticists take a holistic approach which would include a general developmental and paediatric history. Nonetheless not all areas may be covered and indeed parents may not mention everything as they may think it not relevant to the consultation, especially where time pressure is an issue. Clinicians may not have seen the child for a period of time in which a new feature may have arisen. This may result in clinicians simply not knowing or not being aware of issues in certain areas. This could account for the evidence for a difference in quantity score seen in the immune, dental, cardiac, gastroenterology, renal, vision and respiratory systems. Parents are likely to be able to report more in these areas than the clinical geneticist who may have only seen the child once or twice and may be unaware of some of these issues. This has also been reported elsewhere in a study of caregiver-reported information in 237 individuals with Koolen de Vries syndrome which identified that childhood asthma and recurrent pneumonia are respiratory features seen in 40% of this group and not reported previously in clinically reported cohorts(10).

Unique codes from parents in GenROC included nasal congestion, recurrent respiratory infection , asthma and loud snoring for the respiratory system and gingival bleeding, fragile teeth and delayed and advanced eruption of teeth and tooth abscess for the dental systems. This represents a strength of parent reported data in that it represents lived experience data which is likely to be different in granularity to clinician data(24, 27).

The neurological system was notably different from the rest with evidence of a difference in detail scores along with the large number of unique codes in CRD. It is likely that this is due to the cohort being made up of children with neurodevelopmental disorders and therefore a high proportion of neurological phenotypes. Clinicians are likely to therefore have and provide, the most information about this system given it may be seen as most relevant to the genetic diagnosis. When reviewing the list of unique codes for neurology for the clinicians these are made up of MRI Brain descriptors (such as polymicrogyria or cerebellar atrophy), seizure phenotypes (such as focal motor seizure, absence seizure) and clinical phenotypes (such as ataxia, tremor, dystonia). It is plausible that some of these terms at least may not be known to parents or that parents may provide some but not all of them when responding. It is also possible that some parents may assume that CRD will provide this data for their child and so may not have provided it all. It is interesting however that the genes with the highest similarity scores were *CACNA1A* and *STXBP*1. These genes represent fairly tightly defined neurological phenotypes and may explain this higher consistency compared to the rest.

Comparison of CRD and PRD with published phenotypes in DDG2P for *ANKRD11,DDX3X,KMT2A,DYRK1A* and *ADNP* showed CRD was more similar to DDG2P than PRD. PRD did not increase the similarity if added to CRD(4d). This is perhaps not surprising though given the DDG2P list is curated from the published literature which currently largely consists of clinically reported phenotypes. This difference was clear for these genes that are more syndromic and multi-system in nature as compared to the more tightly defined predominantly neurological phenotype genes *TUBA1A, GRIN2B, CASK, STXBP1* and *CACNA1A* in which we found no evidence for a difference between CRD, PRD or combined when generating similarity scores with DDG2P.

## Conclusions

Parents report a similar amount of data to clinicians, but the content differs and is more likely to represent common childhood phenotypes. Clinician reported data is likely to miss that lived experience granularity but is more likely to contain specific clinical phenotypes eg. MRI findings which may be missed or may not be known to parents. The gold standard would be to include both data sources, but this is unlikely to be the case for most studies due to time, cost and resource implications. These nuances should be considered by researchers who are setting up studies or who may be choosing to use specific datasets and when considering the strengths and limitations of said datasets for future applications. Lived experience data should be included in future publications to ensure fullness of phenotype descriptions.

## Data Availability

All data produced in the present study are available upon reasonable request to the authors

## Author contributions

KJL: Conceptualisation; data curation; formal analysis; investigation; methodology; project administration; visualisation; writing – original draft preparation; writing review and editing

HD: methodology, writing review and editing

GenROC Consortium: Data collection

M.T :Data curation

C.D: Data curation

HVF: Methodology; supervision; Writing-review & editing

C.W: Methodology; formal analysis; supervision; Writing-review & editing

## GenROC consortium

Suzanne Alsters, Ruth Armstrong, Tazeen Ashraf, Meena Balasubramanian, Diana Baralle, Jonathan Berg, Ian Berry, Marta Bertoli, Ishita Bhatnagar, Moira Blyth, Thomas Boddington, Catherine Breen, Helen Brittain, Lisa Bryson, Anna de Burca, Jenny Carmichael, Emma Clement, Cristina Dias, Fleur S. Van Dijk, Alan Donaldson, Andrew Douglas, Jacqueline Eason, Sahar Elkady, Nour Elkhateeb, Fayadh Fauzi, Helen V. Firth, Elaine Fletcher, Andrew E. Fry, Laura Furness, Gabriella Gazdagh, Merrie Gowie, Abigail Green, Asma Hamad, Lizzie Harris, Rachel Harrison, Verity Hartill, Eleanor Hay, Jenny Higgs, Jonathon Hoffman, Simon Holden, Samantha Holman, Daniela Iancu, Rachel Irving, Vani Jain, Rosalyn Jewell, Diana Johnson, Gabriela Jones, Beckie Kaemba, Arveen Kamath, Ayse Nur Kavasoglu, Tabassum Khan, Mira Kharbanda, Sophie King, Alison Kraus, Ajith Kumar, Katherine Lachlan, Neeta Lakhani, Wayne Lam, Anne Lampe, Abigail Lazenbury, Harry Leitch, Helen Leveret, Samuel Liebert, Jessica Maiden, Anirban Majumdar, Alison Male, Ruth McGowan, Holly McHale, Alisdair McNeil, Catherine McWilliam, Jonathan Memish, Radwa Mohamed, Tara Montgomery, Oliver Murch, Fiona Osborne, Michael Parker, Vijayalakshmi Ramakumaran, Thiloka Ratnaike, Ruth Richardson, Lisa Robertson, Alison Ross, Claire Searle, Wofah Selah, Resifina Seyara, Charles Shaw-Smith, Suresh Somarathi, Karen Stals, Charlotte Stanley, Edward Steel, A Stewart, Helen Stewart, Kerra Templeton, Riya Tharakan, Madeline Tooley, Mohamed Wafik, Emma Wakeling, Elizabeth Wall, Amy Watford, Patricia Wells, Louise Wilson, Emily Woods

